# Factors Affecting Access to and Utilisation of Intravenous Iron to Treat Anaemia in Pregnancy in Zomba, Malawi: A Qualitative Study

**DOI:** 10.1101/2025.01.30.25321394

**Authors:** Elisabeth Mamani-Mategula, Hana Sabanovic, Naomi Von-Dinklage, Ebony Verbunt, Khic-Houy Prang, Effie Chipeta, Lucinda Manda-Taylor

## Abstract

Anaemia in pregnancy increases risks for both maternal and neonatal complications, including death, preterm birth, and low birth weight. Iron tablets are the recommended standard treatment but are often poorly tolerated and adhered to. Intravenous (IV) iron offers an effective and practical alternative with faster replenishment of iron stores and fewer side effects. A randomised controlled trial in Malawi evaluated the safety and efficacy of IV iron compared to standard oral iron supplementation for pregnant women with moderate to severe anaemia in the third trimester (REVAMP-TT). Our qualitative study, embedded within the trial, aimed to explore the factors affecting access to and utilisation of IV iron to treat anaemia in pregnancy within the primary healthcare system of Malawi.

**Methodology:** We conducted in-depth interviews (n=16) and focus group discussions (n=3) with pregnant women who participated in the REVAMP-TT trial, those who withdrew, and their husbands and caregivers. All interviews and discussions were audio-recorded, transcribed, and coded in NVivo 12. We iteratively used reflexive thematic analysis to develop the themes mapped across the supply and demand domains of the Patient-Centered Access to Healthcare framework.

**Results:** We identified five key themes under supply-side barriers, including i) lack of transparency in REVAMP-TT trial procedures and processes, ii) lack of continued community sensitisation about IV iron, iii) long distances from home to the health facilities, iv) long waiting times for procedures and IV iron administration, and v) pregnant women non-compliance with appointments. Four demand-side obstacles were highlighted, including i) myths and misconceptions about IV iron, ii) prevailing cultural norms like concealing pregnancy, iii) lack of social and financial support from husbands, and iv) physical discomfort when receiving IV iron. Five facilitators emerged from the supply side, including the i) availability of clear information about anaemia and antenatal care outreach services, ii) pregnant women were not pressured to participate in the REVAMP-TT trial, iii) flexible health facility opening hours and appointment mechanism, iv) perceived effectiveness and benefits of IV iron and v) healthcare providers’ interpersonal quality and skills. Four demand-side enablers included i) health literacy about anaemia, ii) social value and a sense of autonomy, iii) peer support, iv) available social and financial support from family or husband, and iv) caregiver support.

**Conclusion:** In conclusion, our study underscores the potential of IV iron therapy to address anaemia in pregnancy in LMICs like Malawi. Patient-centred approaches, improved health literacy, and strengthening health systems are vital for optimising intervention uptake and ensuring equitable access to antenatal care interventions, ultimately improving the health outcomes for mother and child.

**Contributions to literature:** Our study demonstrates the practical aspects and challenges of implementing an IV iron intervention in a low-resource setting, including how the Malawian healthcare system could be strengthened to effectively deliver IV iron in local healthcare facilities. It provides a blueprint for implementation in similar contexts to integrate this treatment into existing healthcare delivery structures.

Our study offers insights into how cultural norms and beliefs shape health-seeking behaviour and practices, contributing to our understanding of how to tailor health interventions to cultural contexts. Additionally, it highlights facilitators, such as local health facilities, that can improve the uptake of IV iron therapy in rural and underserved areas, showcasing how these approaches can be scaled up to enhance maternal health.

## Introduction

Anaemia in pregnancy continues to be a significant global health challenge, carrying enduring consequences for both the mother and child (1–3). Anaemia is defined as a haemoglobin concentration of less than 11 g/dL, leading to a reduced oxygen-carrying capacity of red blood cells to body tissues (1). Approximately 40% of pregnant women worldwide experience anaemia, with the highest burden in low– and middle-income countries (LMICs) (4). Notably, Sub-Saharan Africa has the highest prevalence of anaemia in pregnancy, exceeding 45%, compared to high-income countries with an (average?) prevalence of 9% (2,5,6). In Malawi, nearly 40% of pregnant women experience anaemia (5,6).

Several interrelated factors contribute to the persisting high rates of anaemia in pregnancy in LMICs. These include limited access to antenatal care services for anaemia treatment, nutrient deficiencies through inadequate diets, infections such as malaria, tuberculosis, HIV, chronic inflammatory diseases such as obesity and kidney disease, and other obstetric conditions (1,7). Symptoms of anaemia in pregnancy are varied and can significantly impact the quality of life during pregnancy. These include fatigue, faintness or dizziness, headache, palmar pallor, shortness of breath, and pale eyes (1,8,9). If left untreated, anaemia poses risks of complications for both mother and child, including increased risk of maternal and child mortality, preterm birth, postpartum anaemia, low birth weight, and poor neurological development resulting in cognitive deficits (7,10,11). Children born to mothers who experienced anaemia in pregnancy are more likely to have delayed development and are at a greater risk of developing anaemia later in life (7,10,11).

The World Health Organisation’s (WHO) Global nutrition targets for 2025 aim to reduce the prevalence of anaemia in pregnant women by half, compared to the baseline of 34% in 2012 (12). The strategy emphasises enhancing consistent and fair access to crucial interventions for preventing, diagnosing, and treating anaemia. This includes better screening, managing inherited blood disorders, and reducing infections, inflammation, and chronic diseases affecting micronutrient levels. These efforts align with Sustainable Development Goal (SDG) 2, aiming to eradicate hunger, specifically SDG Target 2.2, which seeks to eliminate all forms of malnutrition, and SDG 3, promoting good health and well-being (12,13). Due to limited progress in reducing the burden of anaemia, the global nutrition target was extended to 2030, and the WHO developed a new comprehensive framework to target the drivers of anaemia (14). The guidelines recommend starting daily supplements of 120 mg of elemental iron as early as possible throughout pregnancy to meet the increased iron demand to support the developing fetus and expand the mother’s red blood cell mass (15). However, this approach faces many challenges, such as low adherence, gastrointestinal effects, limited stocks, and the slow replenishment of iron stores (16–19).

Intravenous (IV) iron is a potential alternative for treating anaemia in pregnancy when oral iron supplementation is ineffective or poorly tolerated (20). A single IV iron infusion, typically administered in 15-30 minutes, can quickly restore blood iron levels without causing gastrointestinal side effects (21–23). Modern IV iron therapies, such as ferric carboxymaltose offers improved safety profiles, better tolerability, and greater efficacy (24).

Numerous studies have highlighted the widespread use and acceptance of IV iron therapy in high-income countries (HICs). This is attributed to their healthcare systems prioritising patient-centred care, IV iron access, and implementing clear guidelines to support evidence-based practices (22, 25–27). For instance, a report from the Australian Government’s Department of Health revealed a doubling in the number of women of reproductive age receiving IV iron in Australia between 2014 and 2017, indicating a high preference for and acceptability of this treatment modality (26).

Although IV iron is increasingly accepted in HICs and some LMICs, several factors affect its utilisation from both supply and demand perspectives. A systematic review of factors and strategies influencing the implementation of IV iron for anaemia in pregnancy identified various barriers and facilitators (28). Supply-side barriers included the high cost of IV iron, including expenses for the drug, hospital admissions, and required personnel; unfamiliarity with IV iron; inadequate resources such as physical space and poor supply chain; and the complexity of preparing and administering IV iron, which is seen as a time-consuming process which further limits its use (27,29–31). Demand side barriers included a lack of knowledge about IV iron, cultural beliefs and misconceptions associating IV iron with vampirism, concerns that it may harm the unborn child, and lack of access to IV iron (31,32). Facilitators for successful IV iron implementation included the perceived advantage of IV iron over oral iron, conducting educational sessions for stakeholders like healthcare providers and pregnant women, and identifying champions to promote IV iron therapy (28,33).

Given the complex interplay of supply and demand factors influencing the uptake of IV iron therapy in pregnant women with anaemia, it is crucial to explore these barriers and facilitators at the primary health facility level. Additionally, it is important to consider how to improve access to and utilisation of IV iron in the context of implementing a randomised controlled trial in Malawi.

## REVAMP-TT trial

A Randomised controlled trial of the Effect of intraVenous iron on Anaemia in Malawian Pregnant women in the Third Trimester (REVAMP-TT) was conducted. REVAMP-TT trial aimed to determine whether a single dose of ferric carboxylmaltose (FCM) compared to oral iron would effectively treat moderate or severe anaemia and improve maternal and child health outcomes at delivery (34). Pregnant women were given one IV iron infusion over 15-30 minutes and were observed for 30 minutes after infusion. Pregnant women were given IV iron in their third trimester because the need for fetal iron transfer is highest, and delivery (with the risk of blood loss) is imminent (35). The cohort of women and their babies enrolled in the REVAMP-TT trial were followed up until 12 months postpartum to assess the treatment’s longer-term benefits and safety. The REVAMP-TT trial was based at the Training and Research Unit of Excellence Centre at Zomba Central Hospital in Malawi. It was operated across the eight primary healthcare settings in the Zomba district. The implementation of the REVAMP-TT trial involved government healthcare providers to represent real-world settings and to explore the applicability and transferability of the intervention to the local context. The findings of the REVAMP-TT trial have been reported elsewhere (ref Santa’s paper).

## REVAMP-IS

An implementation science (IS) research program known as REVAMP-IS was nested within the REVAMP-TT trial. A protocol paper detailing this research program has been published (34). Briefly, the REVAMP-IS program was structured into three phases designed to generate evidence to promote the use of IV iron therapy to treat anaemia in pregnancy in Malawi’s primary healthcare setting. In phase 1, we interviewed key informants including policymakers and health system managers to identify potential barriers and facilitators to implementing IV iron in the Malawian healthcare system. We then conducted co-design workshops with different stakeholders, including healthcare providers and pregnant women, to develop implementation strategies to address the barriers identified and to enhance the uptake and delivery of the IV iron intervention (37). In phase two, we co-developed and co-created educational posters and wall charts highlighting the signs and symptoms of anaemia, the consequences of untreated anaemia, and actionable steps pregnant women can take to prevent or manage their condition. These were for health education and used during the implementation of the REVAMP-TT trial (37). Findings from phases one and two have been previously published (36). In this paper, we present the key findings from the phase three evaluation, which investigated the acceptability and feasibility of IV iron therapy. We applied the Patient-Centered Access to Healthcare (PCAH) Framework to guide this evaluation, emphasising the interplay between demand– and supply-side factors in shaping overall healthcare access and experiences. We explored four domains from the demand side and four from the supply side. We did not include the affordability and ability to pay domains in this analysis as these will be reported in a separate health economics cost evaluation paper. Table 1 outlines the definitions of the PCAH dimensions described by Levesque (38).

## Aim

In this paper, we investigated the factors affecting access to and utilisation of IV iron to treat anaemia in pregnancy in Malawi’s primary healthcare system. From the end-users’ perspective, we investigated the supply and demand barriers and facilitators of access to and utilisation of IV iron. Utilisation is defined as whether people know they need care, want to obtain care, and can access care (39,40). Access is defined as reaching and receiving healthcare promptly to achieve the best possible health outcome (40). Access is examined by the characteristics of individuals, households, social and physical environments, and the attributes of health systems, organisations, and providers (38).

## Methodology

This qualitative study involved in-depth interviews (IDIs) with pregnant women who received IV iron from the eight study sites. We also conducted focus group discussions (FGDs) with pregnant women and their caregivers, such as husbands, mothers, and mothers-in-law. We used the same interview tool guide for the IDIs and FGDs, which was developed based on the demand– and supply-side factors from the PCAH framework (41). From the demand side, we aimed to explore pregnant women’s ability to perceive their healthcare needs, seek healthcare, reach healthcare services, and engage with the healthcare system in accessing IV iron. On the supply side, we focused on understanding the healthcare system’s approachability, acceptability, availability and accommodation, and appropriateness in delivering IV iron therapy. Our reporting adhered to the Consolidated Criteria for Reporting Qualitative Research (COREQ) guidelines (appendix 1) (42).

## Study setting

We conducted our phenomenological qualitative study in Zomba, a district in southeast Malawi. Four primary healthcare facilities were located in urban areas (City Clinic, Sadzi, Matawale, and Naisi) and four in rural areas (Domasi Rural Hospital, Bimbi, Likangala, and Lambulira). All participating public healthcare facilities offer free services, including antenatal care, where pregnant women can access labour, delivery, and postnatal care.

## Participant sampling

### i. In-depth interviews

For the IDIs, we (EMM) randomly selected 42 pregnant women (10%) from a total of 420 listed in the REVAMP-TT trial register, with the sample stratified by sites to ensure that two participants were chosen from each of the eight sites. The trial coordinator provided their identification numbers and contact information, and we contacted them via phone calls and home visits to invite them to participate. We included pregnant women who had received IV iron treatment and were over 18 years old. To ensure representation from each site in case some declined, we randomly selected 19 participants from the 42. Of these, 16 agreed to participate, while three declined because they had withdrawn from the REVAMP-TT trial.

## Focus group discussions

After conducting the IDIs, we (EMM, GK) organised three FGDs to triangulate our data collection methods. We invited pregnant women and their caregivers, such as husbands, mothers, and mothers-in-law, as their views on IV iron use are essential due to their role as key decision-makers in the family. The FGDs fostered a collaborative dialogue, providing a deeper understanding of the social and cultural factors influencing IV iron use and ensuring a wide range of perspectives for more comprehensive feedback. This approach enhanced the validity of our findings and helped minimise research biases.

From the original 10% participants list, we invited the remaining 23 pregnant women, excluding those who had already participated in the IDIs or declined to participate. Nineteen women agreed to participate, while four declined. The FGDs were organised as follows:

1. **First FGD (n=10)**: Comprised pregnant women currently enrolled and actively participating in the REVAMP-TT trial.
2. **Second FGD (n=9)**: Involved pregnant women who had withdrawn from the REVAMP-TT trial (n=3), along with their husbands (n=3) and other pregnant women still participating in the trial (n=3).
3. **Third FGD (n=12)**: Involved pregnant women who were still enrolled and actively participating in the REVAMP-TT trial (n=6), their husbands (n=3), and caregivers (n=3).

## Data collection

Using an interview guide, we (EMM, GK, WS) conducted one-on-one IDIs with pregnant women at the health facilities where they accessed IV iron. IDIs were conducted between September 2022 and February 2023. The IDIs lasted approximately 45 minutes to 1 hour. After conducting the IDIs, we held the FGDs during the same period. Two facilitators were responsible for leading the FGDs: one (EMM) facilitated the discussion, while the other (GK) took notes and made recordings. Each FGD lasted between 1 hour 30 minutes to 2 hours. We invited the participants to a health facility near their homes. We used a designated private room at the health facility for the interviews to ensure a calm and private environment for them to express their views. An interview guide with questions and prompts was used to balance flexibility in the conversation with adherence to the structured framework for data collection. As preferred by the participants, all FGDs and IDIs were conducted in the local language, Chichewa. All interviews were recorded on a digital recorder and we made field notes throughout the data collection process which were used for debriefing sessions. All participants received US$10 in compensation, following the National Health Sciences Research Committee in Malawi’s recommendations for human subjects participating in research (43).

## Data analysis

Two research assistants (WS, GK) transcribed all the interviews verbatim into English, and EMM crosschecked the transcripts for data curation and accuracy. During data analysis, we employed Reflexive Thematic Analysis (RTA). We first generated themes inductively from the data without focusing on a structure or framework to enable in-depth and flexible engagement with the data. We (EMM, HS) independently familiarised ourselves with the dataset to understand its depth and rapidly generated initial themes, which led to the development a preliminary codebook with descriptions and definitions of the themes. We refined the initial codebook through discussions, leading to a consensus on the final version of the codebook. EMM coded all the transcripts to identify detailed codes and new codes iteratively until no new themes or insights emerged from the data. We used NVivo 12 software to organise, manage, and facilitate the comparison and synthesis of the data, ensuring a systematic and comprehensive analysis process. We then reviewed, refined, and clustered the emerging themes and deductively mapped them to the PCAH framework to evaluate and ensure coherence and alignment with the study objectives and outcomes. This combined inductive and deductive approach ensured the analysis was data-driven and grounded in a structured framework.

## Researcher’s Positionality and Reflexivity

This paper reflects a perspective shaped by the researcher’s background in health research and social sciences, with experience implementing health interventions in LMICs, particularly Malawi. The approach is rooted in community-driven, participatory methodologies and guided by principles of advocacy, social justice, equity, and the integration of evidence-based practices into routine care.

This positionality influenced the study design by prioritising methods that amplify the voices of pregnant women and their family members, who are key decision-makers in the family. During data collection, care was taken to foster an inclusive and respectful environment, ensuring that diverse perspectives were actively sought and valued. The participatory approach employed in this study reflects a commitment to co-creating context-specific solutions that address both individual and health system barriers to healthcare access.

Reflexivity was critical in interpreting the findings. Awareness of the researcher’s positionality informed efforts to critically examine biases and ensure that interpretations remained grounded in participants’ realities. The emphasis on equity and inclusivity shaped how data were framed and presented, highlighting barriers, facilitators and strategies to address them.

The commitment to fostering health-promoting practices underscored the importance of considering the socio-cultural context of the communities involved when implementing the intervention. Their perspectives guided the interpretation of findings and the development of recommendations.

## Transparency and credibility

The first author (EMM) piloted the interview guide with two pregnant women participating in the REVAMP-TT trial and who were not part of this qualitative study. Feedback from the pilot informed reflections on the data collection process, leading to refinements of the tool in collaboration with two research assistants (GK, WS) experienced in qualitative research.

Adjustments included rephrasing questions that participants found difficult to understand and adding probes to gain deeper insights into participants’ perspectives. After refining the data collection tools, the team conducted daily debriefing meetings during the 6 months of data collection. These meetings allowed the team to review questions, address areas of concern, and implement necessary revisions or amendments to improve clarity and ensure comprehensive data collection. Data collection methods, including FGDs, IDIs, facilitated data triangulation and validation, enhancing the study’s credibility.

## Consent and ethics clearance

We received ethics approval from the University of Malawi’s College of Medicine Research and Ethics Committee (P.08/20/3114). Permission to collect data from the health facilities was obtained from the Zomba District Health Office (DHO) research committee. We provided the participants with written informed consent forms for their signature or thumbprint for voluntarily agreeing to participate in our study. We gave all participants a copy of their written consent form and reimbursed them for their time and travel expenses per Malawi’s national participant remuneration guidelines (21).

## Results

We conducted 16 IDIs involving two participants per site and three FGDs among 31 participants recruited from the eight sites. Table 2 summarises the total number of participants who consented and were interviewed for IDIs and FGDs.

**Table 1:**
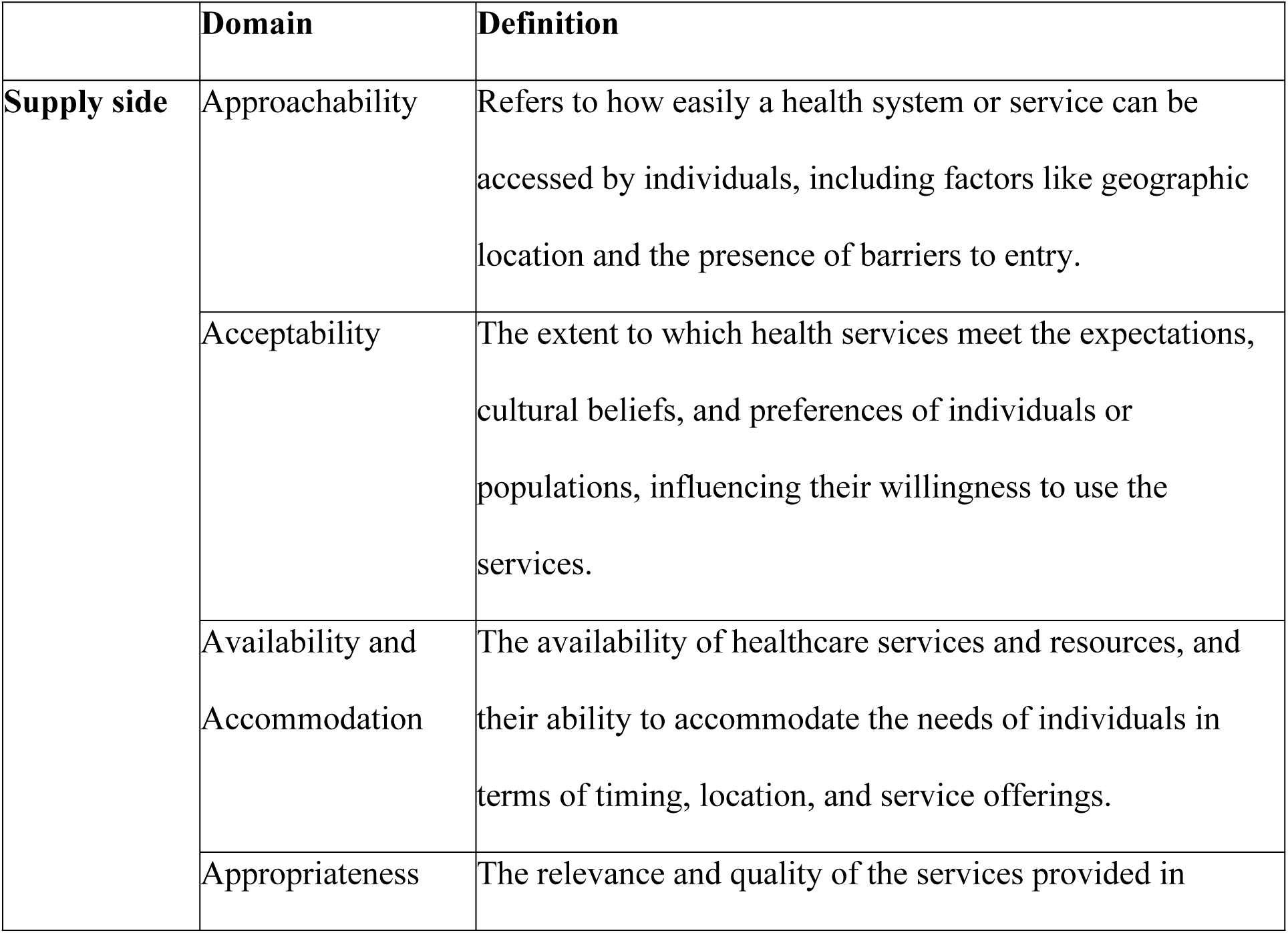
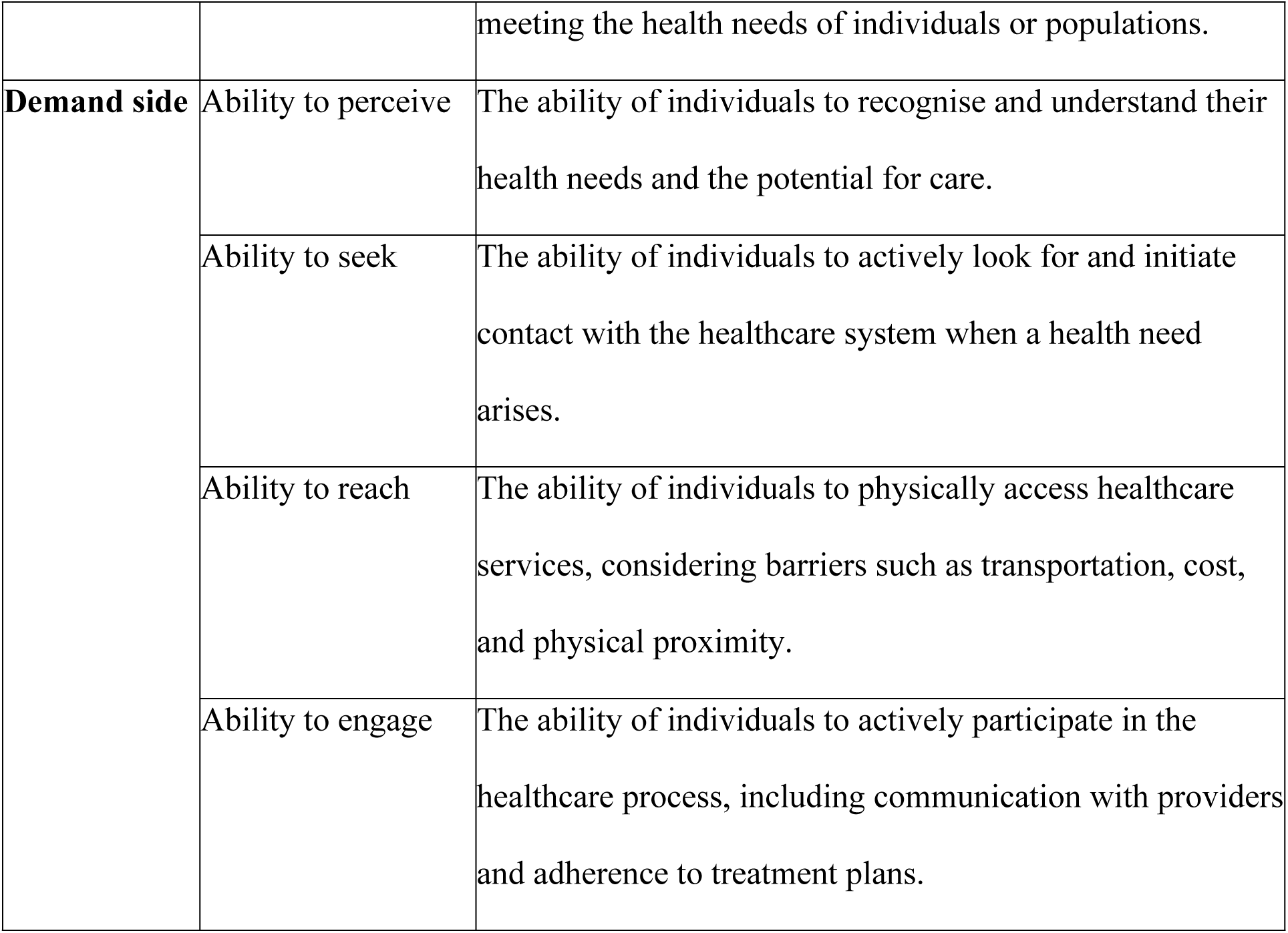
Definitions of the supply and demand domains of the PCAH framework.

|  | **Domain** | **Definition** |
| --- | --- | --- |
| **Supply side** | Approachability | Refers to how easily a health system or service can be accessed by individuals, including factors like geographic location and the presence of barriers to entry. |
|  | Acceptability | The extent to which health services meet the expectations, cultural beliefs, and preferences of individuals or populations, influencing their willingness to use the services. |
|  | Availability and Accommodation | The availability of healthcare services and resources, and their ability to accommodate the needs of individuals in terms of timing, location, and service offerings. |
|  | Appropriateness | The relevance and quality of the services provided in |
|  |  | meeting the health needs of individuals or populations. |
| **Demand side** | Ability to perceive | The ability of individuals to recognise and understand their health needs and the potential for care. |
|  | Ability to seek | The ability of individuals to actively look for and initiate contact with the healthcare system when a health need arises. |
|  | Ability to reach | The ability of individuals to physically access healthcare services, considering barriers such as transportation, cost, and physical proximity. |
|  | Ability to engage | The ability of individuals to actively participate in the healthcare process, including communication with providers and adherence to treatment plans. |

**Table 2:** Summary of participants recruited.

| Participant group |  | Number of participants |  |  |  |
| --- | --- | --- | --- | --- | --- |
|  |  | IDIs | FGD1 | FGD2 | FGD3 |
| 1 | Pregnant women who received IV iron | 16 | 10 | 3 | 6 |
| 2 | Pregnant women who received IV iron but withdrew from REVAMP-TT trial |  |  | 3 |  |
| 3 | Husbands to pregnant women who withdrew from REVAMP-TT trial |  |  | 3 |  |
| 4 | Husbands to pregnant women who received IV iron |  |  |  | 3 |
| 5 | Mothers and mothers-in-law of the pregnant women who received IV iron |  |  |  | 3 |
| Total number of participants |  | 16 | 10 | 9 | 12 |

Table 3 presents the demographic characteristics of participants, including pregnant women, their husbands, and mothers or mothers-in-law who attended the in-depth interviews (IDIs) and the focus group discussions (FGDs).

**Table 3:**
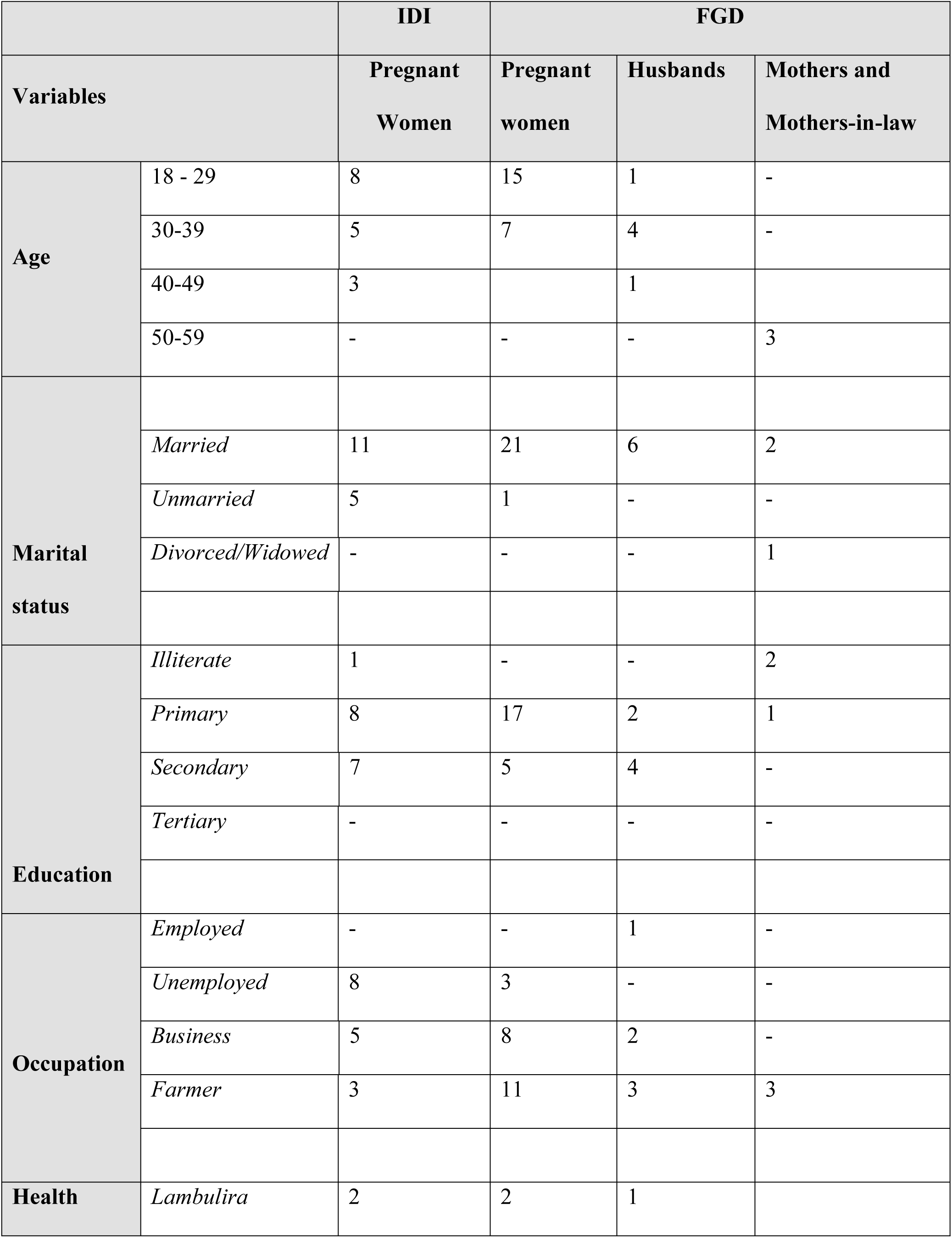
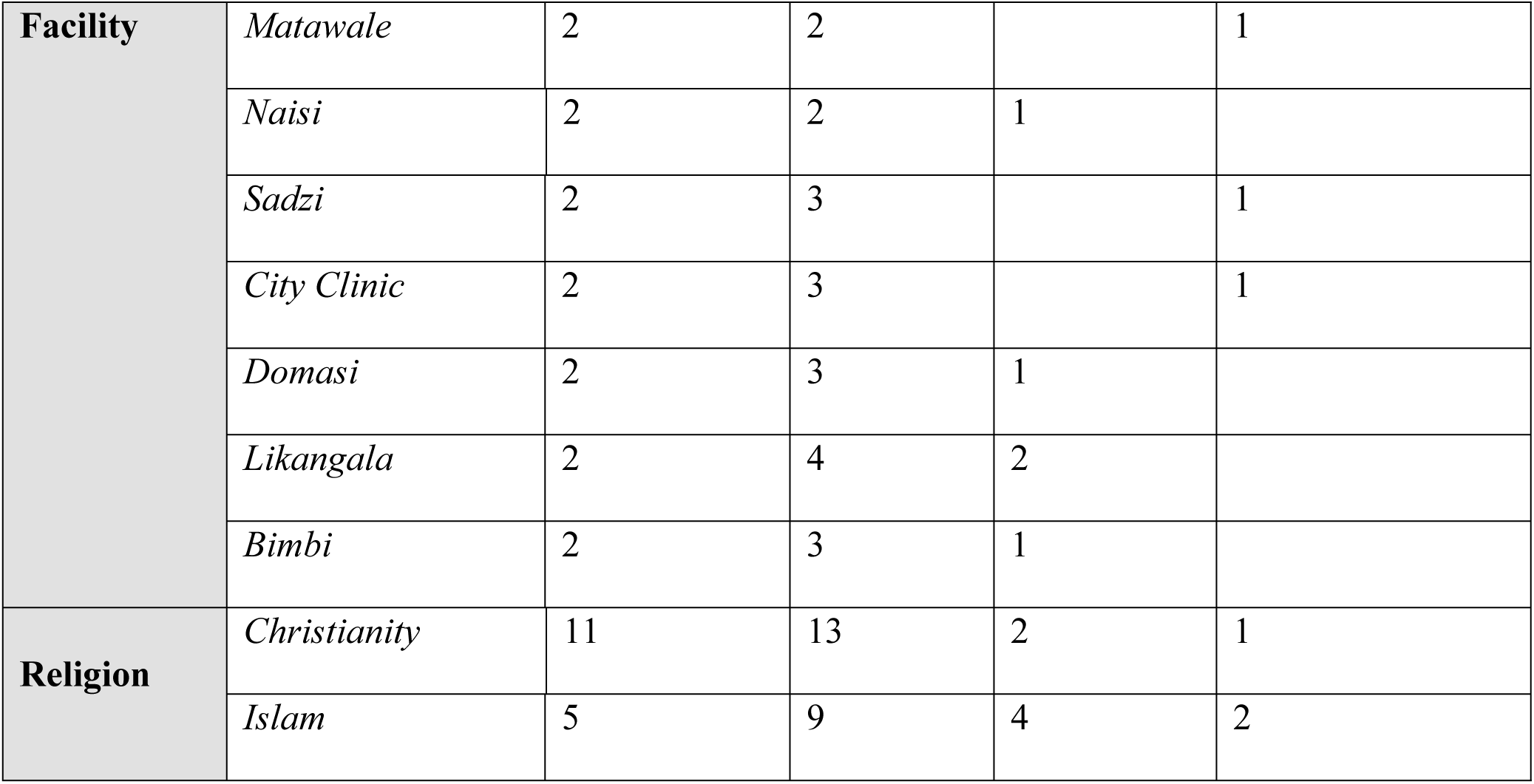
Participant demographic characteristics.

In our study, we explored the demand– and supply-side barriers and facilitators affecting the access to and utilisation of IV iron to treat anaemia in pregnancy in the primary healthcare system of Malawi. We present our findings mapped to the PCAH framework in Table 4.

**Table 4:** A summary of the findings highlighting the supply and demand barriers and facilitators.

| **Category** | **Barriers** | **Facilitators** |
| --- | --- | --- |
| **Supply-Side** | Approachability<br>– Lack of transparency specific to REVAMP-TT trial procedures and processes.<br>– Lack of continued community sensitisation about IV iron. | Approachability - Availability of clear information about anaemia and antenatal care outreach services. |
|  |  | Acceptability – Pregnant women were not pressured to participate in the REVAMP-TT trial. |
|  | Availability and accommodation<br>– Long distance from home to the health facilities.<br>- Long waiting times for procedure and IV iron administration. | Availability and accommodation - Flexible opening hours and appointments mechanism. |
|  | Appropriateness – Pregnant women’s non-compliance with health facility appointments. | Appropriateness<br>– Perceived effectiveness and benefits of IV iron.<br>– Healthcare providers’ interpersonal quality and skills. |
| **Demand-Side** | Ability to perceive - Health beliefs, myths and misconceptions about IV iron. | Ability to perceive - Health literacy, pregnant women expressing awareness of |
|  |  | the signs, symptoms and consequences of anaemia. |
|  | Ability to seek - Prevailing cultural norms such as concealing pregnancy. | Ability to seek<br>Social values and a sense of autonomy influenced informed decision-making.<br>- Peer support in sharing information and experiences informed decision-making. |
|  | Ability to reach - Mobility concerns and a lack of social and financial support from husbands. | Ability to reach<br>– Available social and financial support from family or husband such as assistance with transportation to health facility and provision of food. |
|  | Ability to engage – Pregnant women experienced physical discomfort when receiving IV iron. | Ability to engage - Caregiver support; emotional and practical support from husband or mother/mother-in-law when receiving IV iron. |

From the supply side, we assessed the domains of approachability, acceptability, availability accommodation, and appropriateness. From the demand side, we assessed the domains of ability to perceive, ability to seek, ability to reach, and ability to engage. All the results presented here are from the end-users’ perspective and their perceptions and experiences of supply and demand side barriers and facilitators.

## Supply-side barriers and facilitators to IV Iron Utilisation

### 1. Supply-side barriers

We identified five key themes across three domains of supply-side barriers that could hinder the use of IV iron for treating anaemia in pregnancy. These stemmed from the domains of i) approachability – lack of transparency specific to REVAMP-TT trial procedures and processes and ii) lack of continued community sensitisation about IV iron iii) availability and accommodation – long distances from home to the health facility, iv) long waiting times for procedure and IV iron administration; and v) appropriateness – pregnant women non-compliance with appointments in health facility.

#### i. Approachability – Lack of transparency specific to REVAMP-TT trial processes and procedures

Our study revealed specific barriers to implementing the REVAMP-TT trial, which would be unlikely to occur in primary care as there would be no need for continued sample collection and follow-up visits. Many pregnant women reported that there was a lack of transparency regarding blood sample collection and its purpose during the REVAMP-TT trial implementation. They indicated a need for further explanation, particularly about the necessity of ongoing blood sample collection and follow-up visits even after childbirth. Some pregnant women reported withdrawing from the trial, believing that since their anaemia had been resolved and they had delivered healthy babies, there was no longer a need for continued sample collection.

> *“I felt better after receiving the IV iron, and my body was normally functioning even after childbirth. Therefore, I felt there was no need for a continued blood sample collection and several follow-up visits.” (IDI, Pregnant woman#2)*

Some husbands of pregnant women who had withdrawn from the REVAMP-TT trial also reported that insufficient information about the trial’s processes and procedures influenced their wives’ decisions to discontinue participation.

> *“When she [wife] came home from the follow-up visit, she told me she was no longer interested in participating in the trial [REVAMP-TT trial]. She felt the more she kept going for follow-up visits, the more she was vulnerable to the speculations of the trial that it was about satanism. Since I had no idea about the trial, I let her withdraw. After all, it is her life.” (FGD3 Husband#4)*

A few pregnant women reported that participation in the REVAMP-TT trial was linked to increased cesarean deliveries because women were required to deliver at a central hospital, raising fears, confusion, and distrust therby prompting some to withdraw before delivery.

> *“When I was told that I would have to deliver at the Zomba Central Hospital, I was afraid because here in the community, once we hear that a pregnant woman has been referred to the central hospital, it is likely that there are complications and that she is going “kumpeni” [referring to cesarean section].” (IDI, Pregnant woman#10)*

Many pregnant women were concerned about collecting blood samples from their newborns and collection of “*nsengwa*,” known as placenta tissue, due to its association with cultural rituals.

> *“It is ok when all that [sample collection] is done on me, but I did not feel comfortable with the idea of constantly poking my child every time we come for follow-up as young as she is, collecting blood samples from her without showing us the results. So, I stopped participating in the trial and did not tell anyone. They followed me home but couldn’t find me.” (FGD2, Pregnant woman#1)*

Moreover, some pregnant women raised concerns about the purpose and functionality of the Magnetic Resonance Imaging (MRI) machine. The MRI was used for assessing the cognitive effects of neurological and developmental outcomes of babies born to mothers who had received IV iron. They reported they had never seen the MRI machine and wondered how it functions on their babies while they are asleep. This contributed to their fears and reluctance to continue participating in the REVAMP-TT trial.

> *“They insist that we bring our babies to the Magnetic Resonance Imaging (MRI) machine while they are asleep. They lay our babies inside the machine and attach all the necessary monitoring sensors. As scary as it is, we begin to think, what if they die in there, and why would they need to go in there while asleep?” (FGD1, Pregnant woman#4)*

#### ii. Approachability – Lack of continued community sensitisation about IV iron

Many pregnant women who received IV iron reported first learning about the intervention at the health facility, which caught them off guard and made it difficult for them to accept the treatment immediately. They expressed the lack of continued awareness in their communities, preventing them from acquiring knowledge about IV iron in advance. Some pregnant women and their husbands suggested using platforms like radio, posters, market days, and community-organised campaigns to enhance their knowledge and encourage the use of IV iron therapy.

> *“Of course, you said that you have already gone to do community awareness. But I will advise that you should continually go to the communities and engage with the local leaders to spread the information about the IV iron intervention because women are still getting pregnant day by day, and they may not be aware of the IV iron intervention; you can also use radio programs as they do on Mzati radio.” (FGD2, Husband #3)*

#### iii. Availability and accommodation – Long distance from home to the health facility

While many pregnant women and their caregivers described the antenatal care services as available and accommodating, they identified long distances to the health facility as a barrier to accessing them, including screening for anaemia and receiving IV iron treatment. On average, many pregnant women reported traveling two hours on foot to access the services. This geographical barrier resulted in some pregnant women missing their appointments, follow-up visits and not returning on the scheduled day for IV iron administration.

> *“For example, I live very far away, and I find it so hard to return to the health facility now and again in this condition on foot. If they followed us in our communities, it could have been better. (FGD#3, Pregnant woman#2)*

#### iv. Availability and accommodation – Long waiting times for procedures and IV iron administration

A significant challenge identified by many pregnant women was the prolonged waiting times at the health facilities, which hindered their timely access to care. Pregnant women reported spending up to six hours undergoing anaemia screening and receiving IV iron therapy. These extended waiting periods, exacerbated by long queues, were a considerable burden for them.

> *“The waiting time was indeed long. I came at 8 o’clock, and I was going home around 2 o’clock. On top of that, I was starving because I had not eaten in the morning. The process was long because, before the treatment, the nurses gave information about the IV iron intervention and allowed us to ask questions.” (IDI, Pregnant woman#3)*

#### v. Appropriateness – Pregnant women’s non-compliance with appointments

Many pregnant women reported that non-compliance with antenatal care appointments is a barrier to the timely access of IV iron intervention. Many pregnant women reported delaying their first antenatal care visit, which limited opportunities for early diagnosis and treatment of anaemia. Participants perceived the nine-month pregnancy period as lengthy. They found frequent appointments and follow-ups time-consuming, resulting in reporting for antenatal care when they were close to delivery. They noted that competing demands, such as household responsibilities and childcare, impeded their ability to attend regular antenatal care appointments.

> *“Sometimes you may think your pregnancy is still settling, so you decide to wait a bit. Meanwhile, time is passing. When you reach six months, you start feeling tired and thinking you should rest and stay home while you also have to take care of your other children. That’s how we miss it [the screening and treatment for anaemia].” (IDI, Pregnant Woman #14)*

### 2. Supply-side facilitators

We identified five themes across the four supply-side domains of the PCAH framework that facilitate the use of IV iron to treat anaemia in pregnancy. These included: i) approachability – availability of clear information about IV iron and anaemia and antenatal care outreach services; ii) acceptability – pregnant women were not pressured to participate in the REVAMP-TT trial; iii) availability and accommodation – flexible opening hours and appointments mechanism; and iv) appropriateness – perceived effectiveness and benefits of IV iron and v) appropriateness-healthcare providers’ interpersonal quality and skills.

#### i. Approachability – Availability of clear information about anaemia and antenatal care outreach services

Many pregnant women reported that clear information and explanations about IV iron treatment from healthcare providers and educational materials on anaemia in pregnancy created under REVAMP-IS during the REVAMP-TT trial significantly improved their understanding of anaemia and supported informed decision-making about IV iron therapy. Pregnant women indicated that these materials simplified their understanding of anaemia-related challenges during pregnancy and encouraged them to receive IV iron.

> *“Whenever I attend an antenatal care visit, the nurse provides all the necessary information and explains what we should expect. She educates us about the dangers of anaemia, how it can affect unborn babies, and the importance of treating it.”* (IDI, Pregnant woman#12)

In addition, many pregnant women and their caregivers stressed that delivering IV iron therapy as part of the antenatal care outreach activities at health facilities enhanced their trust in healthcare services. Offering IV iron alongside existing antenatal care services reinforced their confidence in the overall quality and reliability of the healthcare system.

> *“We trust the services that are offered by the nurses we already know at the health facility; they are the ones who help us here in the communities, and there is no way researchers would come directly to us without passing through them.”* (FGD 2, Mother-in #4)

#### ii. Acceptability – Pregnant women were not pressured to participate in the REVAMP-TT trial

Pregnant women highlighted that they were not forced to participate in the REVAMP-TT trial. They reported that informed consent was consistently sought before administering IV iron, fostering a supportive and respectful environment that facilitated acceptance of the intervention. They also applauded the professional communication between them and the healthcare providers as respectful and adequate, accounting for the patient’s background, literacy level, and emotional state differences.

> *“It was easy for me to accept because I was briefed, and they [nurses] gave me a form with all the information about the trial. It explained some of the side effects of the treatment, like feeling weak. So, the nurse assured me that if I ever experience such, I should not be worried and assured to look after me if anything goes wrong.” (IDI, Pregnant woman#4)*

#### iii. Availability and accommodation – Flexible opening hours and appointments

Pregnant women expressed appreciation for the availability and accessibility of the trial facilities, which operated throughout the day on designated antenatal care service days. They reported that they were free to discuss with healthcare providers on scheduled follow-up appointments to ensure continuity of care. Additionally, pregnant women were encouraged to consult their husbands before IV iron administration, with the option to return the next day for the procedure. This flexible approach respected the role of family involvement and helped alleviate stress and anxiety for the pregnant women.

> *“When I found out about my condition, I was filled with worry because I hear a lot of bad stories regarding this [IV iron] treatment. I hear that the health facility takes much blood and sells it. So, I went home, and I told my husband. At that time, I had swollen feet. So, my husband was like, “You have swollen feet, and you look pale because you have low blood levels”, so he escorted me the next day to the health facility where I received the [IV iron] treatment. Then I noticed that everything that people were talking about, I never experienced them.” (FGD1, Pregnant woman#4)*

#### iv. Appropriateness – Perceived effectiveness and benefits of IV iron

Pregnant women who received IV iron reported that they experienced significant improvements in their health, supporting the intervention’s effectiveness. Many pregnant women reported reduced anaemia symptoms, such as headache, dizziness, and fatigue, within two to three days of the treatment. They also expressed satisfaction with the single-dose administration of IV iron, highlighting the convenience of avoiding daily oral iron supplements and frequent visits to the health facility. Additionally, they appreciated that the treatment eliminated the gastrointestinal side effects, such as nausea, commonly associated with oral iron, making it a more reliable and comfortable option for managing anaemia in pregnancy.

> *“Yes, I approve of the IV iron intervention. After receiving the IV iron treatment, I noticed a significant change; unlike the [oral iron] tablet one, I was feeling nauseous and dizzy and not seeing any change. In addition, IV iron is taken once, which I like, unlike the iron tablet, which you are supposed to take daily.” (IDI, Pregnant woman#1)*

#### v. Appropriateness – Healthcare providers interpersonal qualities and skills

Many pregnant women expressed that they found the IV iron intervention appropriately delivered. They appreciated the dedication of the skilled healthcare providers in providing the service and expressed no concerns about the treatment given or being treated by either male or female healthcare providers. Pregnant women emphasised that the intervention was well-suited for their needs, contributing to improvements in their health and overall well-being.

> *“There was nothing scary, as people used to say. The nurses were very friendly and allowed me to ask any questions. They did not even force me to participate. It was my own choice after learning of my condition and the possible IV iron treatment.” (IDI, Pregnant woman#8)*

## B. Demand-side barriers and facilitators to IV Iron Utilisation

### 1. Demand-side barriers

Within the four domains of the PCAH framework, we identified four key themes that represent the barriers to utilising IV iron for managing anaemia in pregnancy, specifically from the demand side. These included i) the ability to perceive – health beliefs, myths, and misconceptions about IV iron, ii) the ability to seek – prevailing cultural norms such as concealing pregnancy, iii) the ability to reach – mobility concerns and a lack of social and financial support from husband, iv) the ability to engage-– physical discomfort when receiving IV iron.

#### i. and ii. Ability to perceive and ability to seek – Myths and misconceptions of IV iron and impact of cultural norms and beliefs

All participants reported myths and misconceptions about IV iron, significantly impacting the utilisation of IV iron therapy. They highlighted that misinformation was widespread among pregnant women attending antenatal care and within the community. Pregnant women reported that some women disregarded medical advice and circulated false claims that IV iron could cause congenital disabilities, malnutrition, or miscarriage. They also noted that some pregnant women diagnosed with anaemia refused to participate in the trial, preferring to use natural remedies, such as *chidede* (hibiscus), to manage their condition.

> *“Ok, what happens is that when we come to attend the antenatal care, some women in the group start spreading rumours that the IV iron is suspicious, we will give birth to abnormal babies, and we don’t know where they take the blood to so we should not agree to receive it when they diagnose us with anaemia. As a result, people start agreeing to their idea, and others leave.” (FGD3, Pregnant woman#6)*

In addition, many participants agreed that some women delay seeking antenatal care and IV iron treatment. Pregnant women often conceal their pregnancy and wait to confirm the visible progression of their pregnancy before attending health facilities for antenatal care services This hesitation stems from fear of becoming the subject of community gossip and apprehension about potential ill will or harmful intentions from others. Such delays result in late diagnosis of anaemia and missed opportunities for timely IV iron intervention.

> *“We wait until we can visibly confirm the pregnancy. Telling everyone that you’re pregnant not only makes you the talk of the community but also exposes you to others who may wish you bad luck.” (FGD 2 Participant #7, Mother)*

#### iii. Ability to reach – Mobility concerns and lack of social and financial support from husbands

Many pregnant women reported that the physical strain of late pregnancy, coupled with long distances to the health facility, made accessing care challenging. As their pregnancies progressed, walking became difficult, and some chose to delay or forgo visits. In the trial, where IV iron was administered in the third trimester, this mobility issue further hindered the timely access to IV iron.

> *“Walking long distances to the health facility is exhausting, especially when you are heavily pregnant. It’s even harder when your husband isn’t available—you almost wish he could experience the challenges we face so he could understand what we go through.” (FGD1, Pregnant woman#7)*

Many pregnant women reported that financial constraints, particularly the lack of support from their husbands, significantly hindered their access to antenatal care and timely IV iron treatment. They reported they often could not afford the transport costs to the health facility for antenatal care appointments, relying on motorbikes or “kabaza,” the primary means of transport in rural Malawi. They said that when faced with the choice between attending appointments or buying food for their families, many opted to skip the appointment, resulting in missed anaemia screening and IV iron administration opportunities.

> *“I separated from my husband, and there was no one to support me. Even if I tried to reason with him that this child belongs to all of us, he has never spent a single penny on this pregnancy. So, because I struggle to make ends meet, I feel the money I find from casual jobs should cater for my food, instead of antenatal care visits.” (IDI, Pregnant woman#14)*

Furthermore, pregnant women reported that the lack of support from their husbands, either due to work commitments or financial constraints, delayed their healthcare decisions to receive IV iron and further postponed receiving necessary treatment.

> *“Unfortunately, my husband was not available when I attended my antenatal care visit and was diagnosed with anaemia. I had to decline the [IV iron] infusion at that moment because I wanted to discuss it with him first. The child is our responsibility, and if I were to lose the pregnancy due to the medication from the health facility, my husband would hold me solely accountable.” (IDI, Pregnant woman#6)*

#### iv. Ability to engage – Physical discomfort during IV iron administration

Pregnant women also reported experiencing physical discomfort due to the IV iron administration, such as pain during cannulation, headaches lasting at several days, and feelings of hunger due to the lengthy procedures, all affecting their physical comfort

> *“I did not like having the cannula on my arm; it was painful and uncomfortable. The waiting time was also long, and I felt hungry because I had not eaten anything since morning.” (IDI, Pregnant woman#8)*

### 2. Demand-side facilitators

We also identified five key themes from the four demand-side domains of the PCAH framework that facilitate the use of IV iron to treat anaemia in pregnancy. These included i) the ability to perceive – health literacy, ii) the ability to seek – social value and a sense of autonomy, peer support, iii) the ability to reach – available social and financial support from family and husband, and iv) the ability to engage – caregiver support during IV iron administration.

#### i. and ii. Ability to perceive and ability to seek – Increased health literacy and peer support about anaemia led to informed healthcare decision-making

Many pregnant women reported that health literacy played a crucial role in their ability to seek IV iron treatment and make informed decisions. They described how understanding their anaemia diagnosis, its consequences, and the benefits of IV iron empowered them to prioritise healthcare facility interventions over traditional or religious healers.

> *“Because I learned about the dangers of anaemia, I was more scared to say no to the treatment when they told me that I was anaemic. I knew it would benefit my life.” (IDI, Pregnant woman#14)*

Additionally, pregnant women reported that shared experiences with peers who had previously received IV iron also influenced them to make informed healthcare decisions. Pregnant women felt reassured hearing about successful outcomes, such as healthy pregnancies and deliveries. This further increased their confidence in the IV iron treatment.

> *“Oh yes! I did. I heard from a friend explaining that the IV iron was not appropriate. They said that the nurses take blood from pregnant women for other things. But I had a relative who had anaemia and was given IV iron. She told me how her condition had changed and encouraged me to receive the [IV iron] treatment, and I disregarded the rumours.” (IDI, Pregnant woman#7)*

In addition, many pregnant women highlighted that their knowledge of anaemia raised their intrinsic and social value of well-being. They emphasised their responsibility to safeguard their health and their unborn children. They said this sense of autonomy enabled them to make independent decisions to receive IV iron without consulting their husbands and relatives, despite societal myths and fears, demonstrating their commitment to improving their health outcomes.

> *“I considered myself lucky when I became aware of my anaemic condition. Myths and misconceptions could not stop me from prioritising my health. After all, my husband and friends could not find a solution to my problem. So, I did what was best for me.”. (IDI, Pregnant woman#7)*

#### iii. Ability to reach – Social and financial support from family and husband

Pregnant women highlighted the importance of social and familial support in facilitating access to IV iron treatment. Many reported that having a caregiver, such as a mother, mother-in-law, or husband, was instrumental in enabling them to reach the health facilities for antenatal care services. They noted that their husbands would accompany them to antenatal care visits on a motorbike or arrange transportation, such as hiring a “kabaza” (bicycle taxi).

> *“My husband was always available to accompany me for my antenatal care visits, and the day I was diagnosed with anaemia to receive IV iron, I was with him.” (FGD1 Pregnant woman#2)*

Pregnant women also reported that financial assistance, such as husbands’ provision of “yodyera kusikelo” (pocket money), was a key motivator for them to attend antenatal care.

> *“Every time, I look forward to my scheduled visit. I benefit a lot. My husband gives me “yodyera kusikelo” (pocket money), making me a proud wife.” (IDI, Pregnant woman#3)*

#### iv. Ability to engage – Caregivers’ support; emotional and practical support from husband or mother/mother-in-law when receiving IV iron

Pregnant women emphasised the importance of emotional support from caregivers, such as husbands, mothers, or mothers-in-law, in encouraging them to receive IV iron treatment. The reassurance and motivation from these key figures helped them overcome fears and doubts during the process and made them more confident in engaging with the IV iron intervention.

> *“When my wife was found with low blood levels and was experiencing heavy dizziness and headache, I was worried because I knew she was at risk of losing her life and the child. When we were told about the IV iron intervention, I encouraged her to accept the treatment, and I was there during the administration. I encouraged her and assured her she would be fine. All we wanted was for her to feel better.” (FGD2, Husband#3)*

## Discussion

Using the PCAH framework, we identified several demand and supply barriers and facilitators affecting IV iron access and utilisation to treat anaemia in pregnancy. Barriers included a lack of transparency in the REVAMP-TT trial procedures, insufficient community sensitisation about IV iron, long travel distances from home to healthcare facilities, long waiting times to receive treatment, pregnant women’s non-compliance with appointments, myths and misconceptions about IV iron, cultural norms, and inadequate social or financial support from husbands. Facilitators included access to clear information about anaemia and antenatal care services, voluntary participation in the REVAMP-TT trial, flexible health facility opening hours, perceived benefits of IV iron, healthcare providers’ interpersonal skills, better health literacy, social support from family or husbands, and caregiver involvement. We demonstrated how supply and demand domains influence healthcare decisions and deepen our understanding of patients’ needs, preferences, and experiences. This approach underscores the need for health interventions that are more effective, accessible, and responsive to the target population.

## Transparency gaps, structural and socio-cultural barriers: A multi-dimensional challenge to IV iron access and utilisation in pregnancy

### Transparency gaps

We identified barriers related to the REVAMP-TT trial. In particular, we found that the lack of transparency regarding the trial procedures and processes was a critical barrier to approaching and receiving IV iron. Key concerns included the amount of blood samples collected, the purpose of the blood samples, and the rationale behind ongoing blood sample collection with several follow-up visits even after childbirth. The lack of clarity on the processes and procedures created uncertainty, anxiety, and hesitancy, discouraging some pregnant women from seeking treatment and causing others to withdraw altogether. This finding aligns with an overview of reviews conducted on the barriers and facilitators to subjects’ participation in clinical trials, where they also reported a lack of transparency and described the information about the trial as too technical and too complex to be easily understood by participants (44). Another literature review on the primary barriers and facilitators to participation in clinical research confirms that a lack of knowledge and understanding of the clinical processes influences negative attitudes toward accepting and participating in clinical trials (45). This suggests that transparency in healthcare processes is a fundamental aspect of successful clinical trial interventions and a critical factor in building trust and promoting informed decision-making (46). The importance of targeted outreach education and effective open and transparent communication cannot be overstated, as it improves knowledge and addresses the mistrust and fears of the participants (45). When pregnant women perceive healthcare providers as withholding or inadequately explaining information, it undermines their confidence in the system, reducing their willingness to engage with interventions like IV iron therapy and leading to missed opportunities for early intervention, delayed treatment, and poor adherence to recommended care. Increased transparency increases awareness and knowledge, consequently enhancing health-seeking behaviour.

### Structural and socio-cultural barriers

Our findings demonstrated that the social, structural, and social-cultural barriers to IV iron access and utilisation are intertwined. For instance, the absence of continuous targeted community awareness campaigns about IV iron leads to increased cultural norms and social beliefs, such as myths and misconceptions about IV iron and behaviours of concealing a pregnancy. As a result, this creates fear and skepticism in pregnant women’s willingness to use IV iron. Likewise, long distances to the health facilities, lack of transportation, long waiting times, and lack of social support from husbands not only discouraged women from accessing IV iron therapy and missing appointments but also contributed to delays in seeking treatment for anaemia in pregnancy. The findings on the barriers reported in this study mirror the conclusions reported in studies on the obstacles to the use of IV iron amongst pregnant women with anaemia in Nigeria (31), Malawi (30,36), Australia (47). Other studies have reported similar barriers to access and use of antenatal care services (48–52). These interconnected barriers for a pregnant woman living in rural areas of Malawi mean discouragement in timely reaching for healthcare services, especially for IV iron, which can only be administered in a health facility setting. Although access to antenatal care and the use of IV iron could face persistent barriers, pregnancy in low-resource settings represents a rare and critical moment when women engage with the health system (53). This unique window must not be wasted; it is an urgent call to dismantle these barriers and ensure women not only access but fully benefit from life-saving interventions (like IV iron) that can improve maternal and child health outcomes. Addressing socio-cultural perceptions and idioms will be crucial in a pregnant woman’s decision to seek care and adhere to treatment.

### Bridging gaps in access to and utilisation of IV iron: a balanced approach between demand and supply side enablers

Several interconnected domains facilitated access to and utilisation of IV iron. The availability of educational information about anaemia and IV iron services played a pivotal role in enabling pregnant women to recognise their healthcare needs and actively seek care. Their health literacy, the social value ascribed to one’s health, and the degree of autonomy in making healthcare decisions influenced them to accept the intervention amidst the reported cultural and social factors. Malawi has made significant progress in antenatal care coverage, where about 97% of pregnant women access antenatal care (54). This highlights that pregnant women have adequate health literacy and understand the importance of attending antenatal care, prompting them to recognise the importance of IV iron therapy and make informed decisions about their health.

Pregnant women in our study consistently reported that promoting male involvement in antenatal care services would foster their acceptability of IV iron. Literature reveals that traditional notions of men as decision-makers and socio-cultural views on health remain vital as men are considered the primary and key decision-makers in many households. Men still play a crucial role in providing financial support in their homes, highlighting their influence in shaping household dynamics and health decisions (55). A study in Somaliland confirmed that pregnant women who were motivated and accompanied by their husbands to antenatal care were more likely to use antenatal care services than those who were not accompanied (56). By fostering shared responsibility between husband and wife for antenatal care, pregnancy can be reframed not solely as a woman’s reproductive roles and responsibilities but as a collaborative effort, thereby redefining the perception that men and women occupy different spaces over which they have authority. This will ultimately make it easy for pregnant women to decide and reach for healthcare interventions such as IV iron, therefore enhancing maternal and child health outcomes.

The goal of any health intervention is to align seamlessly with the patient’s needs, ensuring its relevance and effectiveness. Our study demonstrates that pregnant women perceived the IV iron intervention as an appropriate and efficacious treatment for anaemia. Moreover, evidence from individuals who had previously benefited from the IV iron intervention proved pivotal in fostering active engagement, as their firsthand experiences provided tangible proof of the treatment’s effectiveness. Women were impressed that, unlike oral iron tablets, IV iron works more rapidly, avoids gastrointestinal side effects, and eliminates the burden of daily medication, making it a more patient-centred approach. These findings confirmed the findings reported in a study conducted in Nigeria, where they found that the perceived comparative advantages of IV iron were critical for acceptability (48). This perceived fit between the intervention and their healthcare needs significantly enhances the acceptability of an intervention, demonstrating the profound impact of compelling evidence in not only validating the appropriateness of the intervention but also ensuring active engagement.

Collectively, these findings signify that addressing factors affecting the uptake of health interventions, such as IV iron, improves the acceptance and use of health interventions for managing anaemia in pregnant women.

## Strengths and Limitations

To our knowledge, this is the first study using the PCAH framework to report on the factors influencing access to and utilisation of IV iron to treat anaemia in pregnant women in Malawi. This provides a robust basis of knowledge for evaluating the entire patient journey experience. Evaluating the IV iron intervention through the end user lens provided a critical and varied assessment of how to increase the utilisation of IV iron to meet the needs of those who should benefit. In this evaluation, we included pregnant women participating in the REVAMP-TT trial and those who withdrew with their husbands and other caregivers from across the eight sites, which enhanced the robustness and applicability of the findings. Additionally, this reduced bias in our findings and allowed us to understand the role of social support during pregnancy. Using IDIs and FGDs helped us understand the full spectrum of participants’ experiences, which can be used to inform the development of more accessible and highly utilised healthcare interventions.

While this paper focuses on the perspectives of pregnant women regarding IV iron utilisation, we did not interview pregnant women who declined to participate in the REVAMP-TT trial or those who received oral iron tablets to understand their perspectives. We did not include the insights from the healthcare providers; however, HS and KP will lead a separate paper that presents their views to allow for a more detailed exploration of their insights. Additionally, although this paper does not delve into the affordability and ability to pay constructs of the PCAH framework, a forthcoming paper will specifically address the cost evaluation, providing a comprehensive assessment of the economic aspects of IV iron implementation. The positive responses to IV iron may have been influenced by the $10 reimbursements given to the participants as part of the trial. It is essential also to evaluate such interventions independently of financial reimbursements.

## Conclusion

Anaemia in pregnancy is a common condition disproportionally affecting women who live in LMICs. Our study examined the demand and supply barriers and facilitators influencing access to and use of intravenous (IV) iron to treat anaemia in pregnancy across eight health facilities in Malawi. The results demonstrate significant potential for IV iron in addressing anaemia, underscoring the importance of patient-centred approaches and effective healthcare delivery. Focusing on improving health literacy and awareness and fostering social support networks would encourage pregnant women to seek essential antenatal care services. Strengthening the health system’s capacity through training for healthcare providers, improving infrastructure and road networks, and enhancing community education is crucial for optimising the utilisation of health interventions, ensuring that they are responsive to the diverse needs of the end users. These efforts will ultimately improve maternal and child health outcomes and promote equity in healthcare access.

## Availability of data and materials

The datasets generated and analysed during the current study could be available from the corresponding author on reasonable request with permission from the College of Medicine Research Ethics Committee

## Data Availability

NA

## Abbreviations

DHO: District Health Office
FGD: Focus group discussion
FCM: Ferric carboxymaltose
HIC: High-income country
IDI: In-depth interview
IV: Intravenous
LMIC: Low-middle-income country
MRI: Magnetic Resonance Imaging
PCAH: Patient-Centred Access to Healthcare
REVAMP-IS: Randomised controlled trial on the effect of intravenous iron on moderate and severe anaemia in pregnant women in the third trimester – Implementation Science
REVAMP-TT: Randomised controlled trial on the effect of intravenous iron on moderate and severe anaemia in pregnant women in the third trimester
RTA: Reflexive thematic analysis
SDG: Sustainable Development Goals

## Acknowledgments

We want to acknowledge the research assistants, Grace Thole-Katola (GTK) and Wakumanya Sibande (WS), who supported the acquisition of the data, the study participants, the dedication of the REVAMP-TT trial staff in Zomba, Malawi, and collaborators at the Walter and Eliza Hall Institute in Melbourne, Australia. We are grateful for the support we continue to receive from the District Health Offices in Zomba, Malawi, and the managerial and nursing staff of the government health facilities where the REVAMP-TT trial was conducted.

## Author Information

### Authors and affiliation

^1^ Department of Health Systems and Policy, Kamuzu University of Health Sciences, Malawi Elisabeth Mamani-Mategula, Effie Chipeta, Lucinda Manda-Taylor.

^2^ Centre for Health Policy, Melbourne School of Population and Global Health, The University of Melbourne, Australia.

Hana Sabanovic, Ebony Verbunt, Khic-Houy Prang.

^3^ Population Health and Immunity division, The Walter and Eliza Hall Institute of Research, Melbourne, Australia.

Naomi Von-Dinklage

## Contributions

EMM, KHP, EC, and LMT designed the study. EMM GTK and WS conducted the in-depth interviews and focus group discussions. EMM, HS, and LMT analysed and interpreted the data. EMM drafted the manuscript, with KHP, EV, HS, NVD, EC, and LMT reviewing and contributing. All authors read and approved the final manuscript.

## Corresponding author

Elisabeth Mamani-Mategula

## Ethics declaration

### Ethics approval and consent

We received ethics approval (P.08/20/3114) from the University of Malawi’s College of Medicine Research and Ethics Committee. All participants signed the informed consent form.

## Consent for publication

Not applicable.

## Appendix 1

**Table 5:**
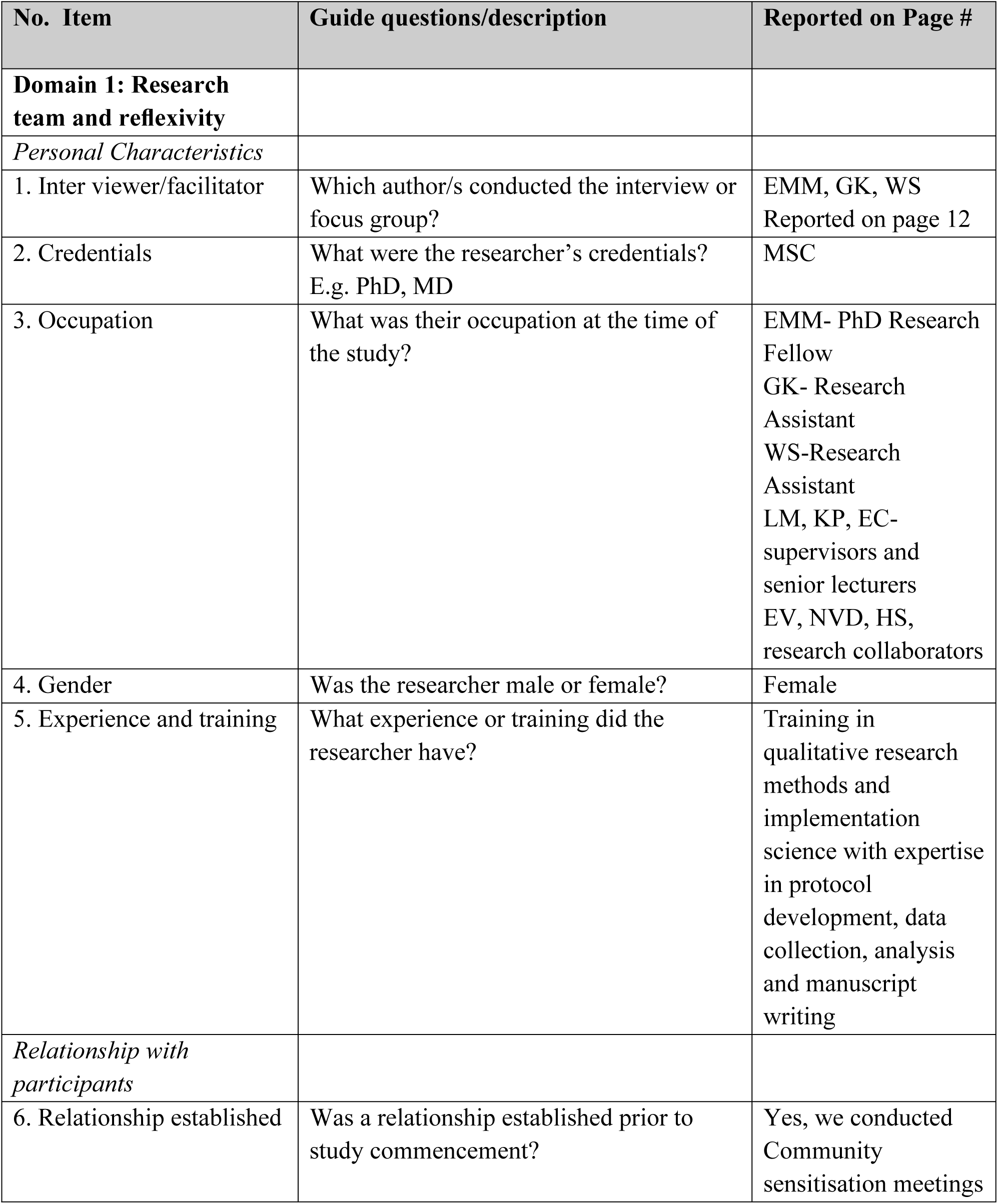
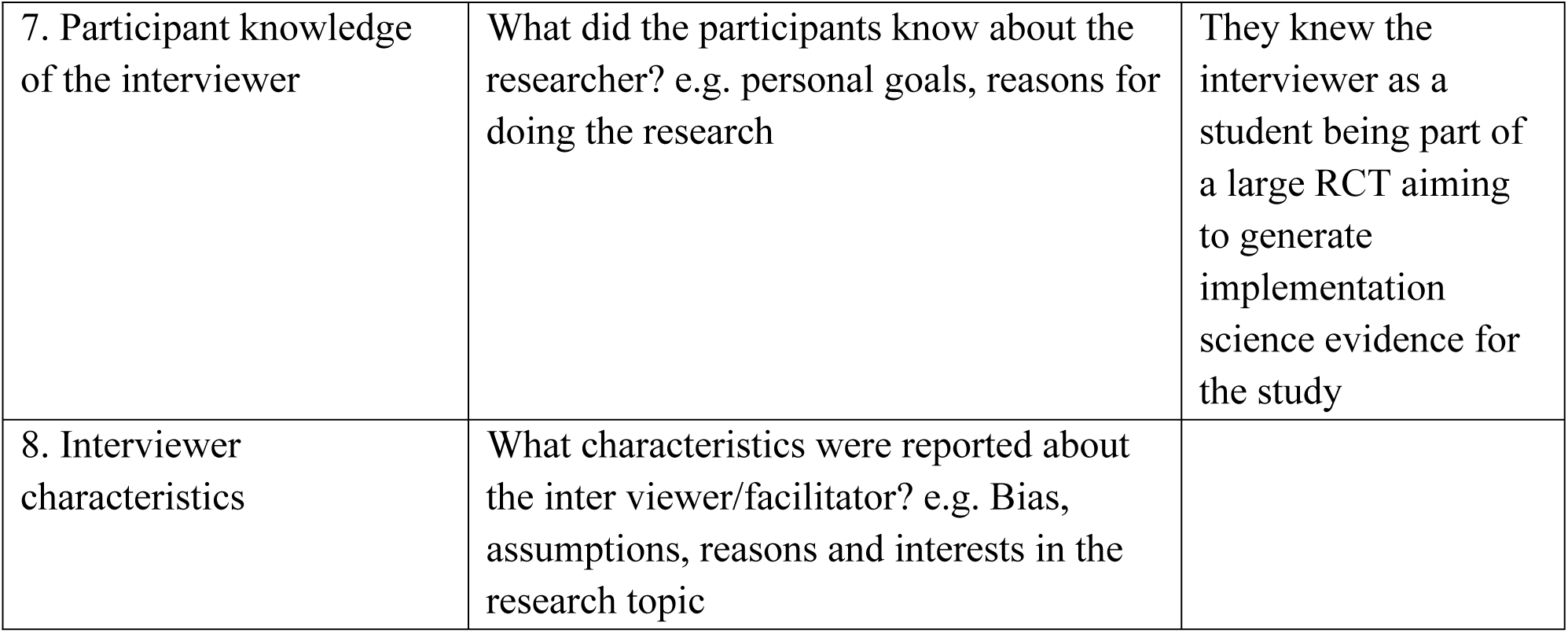
Consolidated criteria for reporting qualitative studies (COREQ): 32-item checklist.

| No. Item | Guide questions/description | Reported on Page # |
| --- | --- | --- |
| **Domain 1: Research team and reflexivity** |  |  |
| *Personal Characteristics* |  |  |
| 1. Inter viewer/facilitator | Which author/s conducted the interview or focus group? | EMM, GK, WS<br>Reported on page 12 |
| 2. Credentials | What were the researcher's credentials?<br>E.g. PhD, MD | MSC |
| 3. Occupation | What was their occupation at the time of the study? | EMM- PhD Research Fellow<br>GK- Research Assistant<br>WS-Research Assistant<br>LM, KP, EC- supervisors and senior lecturers<br>EV, NVD, HS, research collaborators |
| 4. Gender | Was the researcher male or female? | Female |
| 5. Experience and training | What experience or training did the researcher have? | Training in qualitative research methods and implementation science with expertise in protocol development, data collection, analysis and manuscript writing |
| *Relationship with participants* |  |  |
| 6. Relationship established | Was a relationship established prior to study commencement? | Yes, we conducted Community sensitisation meetings |
| 7. Participant knowledge of the interviewer | What did the participants know about the researcher? e.g. personal goals, reasons for doing the research | They knew the interviewer as a student being part of a large RCT aiming to generate implementation science evidence for the study |
| 8. Interviewer characteristics | What characteristics were reported about the interviewer/facilitator? e.g. Bias, assumptions, reasons and interests in the research topic |  |

## References

1. Shand AW, Kidson-Gerber GL. Anaemia in pregnancy: a major global health problem. The Lancet [Internet]. 2023 May 13 [cited 2024 Sep 29];401(10388):1550–1. Available from: http://www.thelancet.com/article/S0140673623003963/fulltext

2. Sujeevani Munasinghe N van den B. Anaemia in pregnancy in Malawi – A Review. Malawi Medical Journal. 2007;18(4):160–74.

3. Gilder ME, Simpson JA, Bancone G, McFarlane L, Shah N, van Aalsburg R, et al. Evaluation of a treatment protocol for anaemia in pregnancy nested in routine antenatal care in a limited-resource setting. Glob Health Action [Internet]. 2019 Jan 1 [cited 2024 Jan 10];12(1). Available from: /pmc/articles/PMC6586122/

4. Anemia — Level 1 impairment | Institute for Health Metrics and Evaluation [Internet]. [cited 2024 Apr 14]. Available from: https://www.healthdata.org/research-analysis/diseases-injuries/factsheets/anemia-level-1-impairment

5. Liyew AM, Tesema GA, Alamneh TS, Worku MG, Teshale AB, Alem AZ, et al. Prevalence and determinants of anaemia among pregnant women in East Africa; A multi-level analysis of recent Demographic and Health Surveys. PLoS One [Internet]. 2021 Apr 1 [cited 2024 Mar 22];16(4):e0250560. Available from: https://journals.plos.org/plosone/article?id=10.1371/journal.pone.0250560

6. Nyarko SH, Boateng ENK, Dickson KS, Adzrago D, Addo IY, Acquah E, et al. Geospatial disparities and predictors of anaemia among pregnant women in Sub-Saharan Africa. BMC Pregnancy Childbirth [Internet]. 2023 Dec 1 [cited 2024 Mar 22];23(1):1–13. Available from: https://bmcpregnancychildbirth.biomedcentral.com/articles/10.1186/s12884-023-06008-3

7. Balcha WF, Eteffa T, Tesfu AA, Alemayehu BA, Chekole FA, Ayenew AA, et al. Factors associated with anaemia among pregnant women attended antenatal care: a health facility-based cross-sectional study. Annals of Medicine and Surgery [Internet]. 2023 May 12 [cited 2024 Mar 12];85(5):1712. Available from: /pmc/articles/PMC10205215/

8. Dwumfour-Asare B, Kwapong M. Anaemia awareness, beliefs and practices among pregnant women: a baseline assessment at Brosankro community in Ghana. Journal of Natural Sciences Research [Internet]. 2014;3(15):1–9. Available from: http://www.iiste.org/Journals/index.php/JNSR/article/view/9709

9. Khalafallah AA, Dennis AE. Iron deficiency anaemia in pregnancy and postpartum: pathophysiology and effect of oral versus intravenous iron therapy. J Pregnancy [Internet]. 2012 [cited 2024 Jul 24];2012. Available from: https://pubmed.ncbi.nlm.nih.gov/22792466/

10. Factors associated with anaemia among pregnant women attended antenatal care: a health facility-based cross-sectional study – PMC [Internet]. [cited 2024 Mar 12]. Available from: https://www.ncbi.nlm.nih.gov/pmc/articles/PMC10205215/

11. Daru J, Zamora J, Fernández-Félix BM, Vogel J, Oladapo OT, Morisaki N, et al. Risk of maternal mortality in women with severe anaemia during pregnancy and postpartum: a multilevel analysis. Lancet Glob Health [Internet]. 2018 May 1 [cited 2024 Oct 24];6(5):e548–54. Available from: https://pubmed.ncbi.nlm.nih.gov/29571592/

12. Global nutrition targets 2025: policy brief series [Internet]. [cited 2024 Mar 12]. Available from: https://www.who.int/publications/i/item/WHO-NMH-NHD-14.2

13. Nations U. 70/1. Transforming our world: the 2030 Agenda for Sustainable Development Transforming our world: the 2030 Agenda for Sustainable Development Preamble. 2030;

14. Accelerating anaemia reduction: A comprehensive framework for action.

15. Antenatal iron supplementation [Internet]. [cited 2024 Mar 12]. Available from: https://www.who.int/data/nutrition/nlis/info/antenatal-iron-supplementation

16. Fouelifack FY, Sama JD, Sone CE. Assessment of adherence to iron supplementation among pregnant women in the Yaounde gynaeco-obstetric and paediatric hospital. Pan African Medical Journal. 2019;34:1–8.

17. Benedict R, Wang W, Mallick L. Examining the Role of Health Facilities in Supporting Iron Folic Acid Supplementation Adherence Among Women in Malawi (OR25-03-19). Curr Dev Nutr. 2019;3(Supplement_1):7200.

18. Ba DM, Ssentongo P, Kjerulff KH, Na M, Liu G, Gao X, et al. Adherence to Iron Supplementation in 22 Sub-Saharan African Countries and Associated Factors among Pregnant Women: A Large Population-Based Study. Curr Dev Nutr. 2019;3(12):1–8.

19. Ba DM, Ssentongo P, Kjerulff KH, Na M, Liu G, Gao X, et al. O R I G I N A L R E S E A R C H Adherence to Iron Supplementation in 22 Sub-Saharan African Countries and Associated Factors among Pregnant Women: A Large Population-Based Study.

20. Gasche C, Berstad A, Befrits R, Beglinger C, Dignass A, Erichsen K, et al. Guidelines on the diagnosis and management of iron deficiency and anaemia in inflammatory bowel diseases. Inflamm Bowel Dis [Internet]. 2007 Dec [cited 2024 Apr 10];13(12):1545–53. Available from: https://pubmed.ncbi.nlm.nih.gov/17985376/

21. Seeho SKM, Morris JM. Intravenous iron use in pregnancy: Ironing out the issues and evidence. Australian and New Zealand Journal of Obstetrics and Gynaecology. 2018;58(2):145–7.

22. World T. Iron-deficiency anaemia in pregnancy and the role of intravenous iron. Contemp Ob Gyn. 2021;(July):20–4.

23. Govindappagari S, Burwick RM. Treatment of Iron Deficiency Anemia in Pregnancy with Intravenous versus Oral Iron: Systematic Review and Meta-analysis. Am J Perinatol [Internet]. 2019 [cited 2024 Apr 14];36(4):366–76. Available from: https://pubmed.ncbi.nlm.nih.gov/30121943/

24. Jimenez K, Kulnigg-Dabsch S, Gasche C. Management of Iron Deficiency Anemia. Gastroenterol Hepatol (N Y). 2015;11.

25. Burn MS, Lundsberg LS, Culhane JF, Partridge C, Son M. Intravenous iron for treatment of iron deficiency anaemia during pregnancy and associated maternal outcomes. The Journal of Maternal-Fetal & Neonatal Medicine [Internet]. 2023 Dec 31 [cited 2024 Feb 7];36(1). Available from: https://www.tandfonline.com/doi/abs/10.1080/14767058.2023.2192855

26. Seeho SKM, Morris JM. Intravenous iron use in pregnancy: Ironing out the issues and evidence. Australian and New Zealand Journal of Obstetrics and Gynaecology [Internet]. 2018 Apr 1 [cited 2023 Nov 23];58(2):145–7. Available from: https://onlinelibrary.wiley.com/doi/full/10.1111/ajo.12794

27. Mayson E, Ampt AJ, Shand AW, Ford JB. Intravenous iron: Barriers and facilitators to its use at nine maternity hospitals in New South Wales, Australia. Australian and New Zealand Journal of Obstetrics and Gynaecology. 2016;56(2):162–72.

28. Verbunt E, Akter S, Manda-Taylor L, Sarker B, Ataide R, Davidson E, et al. Factors and strategies influencing implementation of an intravenous iron intervention for antenatal anaemia: A mixed-methods systematic review. Reproductive, Female and Child Health. 2023 Jun;2(2):59–70.

29. Smith-Wade S, Kidson-Gerber G, Shand A, Grzeskowiak L, Henry A. The use of intravenous iron in pregnancy: for whom and when? A survey of Australian and New Zealand obstetricians. BMC Pregnancy Childbirth. 2020 Dec 1;20(1).

30. Manda-Taylor L, Kufankomwe M, Chatha G, Chipeta E, Mamani-Mategula E, Mwangi MN, et al. Perceptions and experiences of intravenous iron treatment for anaemia in pregnancy in Malawi: a formative qualitative study. Gates Open Res [Internet]. 2022 Nov 8 [cited 2024 Mar 14];6:66. Available from: https://pubmed.ncbi.nlm.nih.gov/38455670/

31. Akinajo OR, Babah OA, Banke-Thomas A, Beňová L, Sam-Agudu NA, Balogun MR, et al. Acceptability of IV iron treatment for iron deficiency anaemia in pregnancy in Nigeria: a qualitative study with pregnant women, domestic decision-makers, and health care providers. Reprod Health. 2024 Dec 1;21(1).

32. Manda-Taylor L, Kufankomwe M, Chatha G, Chipeta E, Mamani-Mategula E, Mwangi MN, et al. Perceptions and experiences of intravenous iron treatment for anaemia in pregnancy in Malawi: a formative qualitative study. Gates Open Res. 2022;6(May):66.

33. Akinajo OR, Babah OA, Banke-Thomas A, Beňová L, Sam-Agudu NA, Balogun MR, et al. Acceptability of IV iron treatment for iron deficiency anaemia in pregnancy in Nigeria: a qualitative study with pregnant women, domestic decision-makers, and health care providers. Reprod Health. 2024 Dec 1;21(1).

34. ACTRN12621001239853. Randomised controlled trial of the Effect of intraVenous iron on Anaemia in Malawian Pregnant women in their third trimester: REVAMP-TT. https://trialsearch.who.int/Trial2.aspx?TrialID=ACTRN12621001239853 [Internet]. [cited 2023 Oct 23]; Available from: https://www.cochranelibrary.com/central/doi/10.1002/central/CN-02327291/full

35. Mwangi MN, Mzembe G, Moya E, Braat S, Harding R, Robberstad B, et al. Protocol for a multicentre, parallel-group, open-label randomised controlled trial comparing ferric carboxymaltose with the standard of care in anaemic Malawian pregnant women: the REVAMP trial. BMJ Open [Internet]. 2021 [cited 2023 Oct 23];11:53288. Available from: http://bmjopen.bmj.com/

36. Mamani-Mategula E, Von-Dinklage N, Sabanovic H, Verbunt E, Prang KH, Chipeta E, et al. Using an experience-based co-design approach to develop strategies for implementing an intravenous iron intervention to treat moderate and severe anaemia in pregnancy in Malawi. Implement Sci Commun [Internet]. 2024 Dec 1 [cited 2024 Dec 2];5(1):1–19. Available from: https://implementationsciencecomms.biomedcentral.com/articles/10.1186/s43058-024-00661-1

37. Mamani-Mategula E, Von-Dinklage N, Sabanovic H, Verbunt E, Prang KH, Chipeta E, et al. Using an experience-based co-design approach to develop strategies for implementing an intravenous iron intervention to treat moderate and severe anemia in pregnancy in Malawi. Implementation Science Communications 2024 5:1 [Internet]. 2024 Nov 15 [cited 2024 Nov 18];5(1):1–19. Available from: https://implementationsciencecomms.biomedcentral.com/articles/10.1186/s43058-024-00661-1

38. Levesque JF, Harris MF, Russell G. Patient-centred access to health care: Conceptualising access at the interface of health systems and populations. Int J Equity Health [Internet]. 2013 Mar 11 [cited 2024 Nov 18];12(1):1–9. Available from: https://equityhealthj.biomedcentral.com/articles/10.1186/1475-9276-12-18

39. Watts AS, Crimmins EM. Populations at special health risk: The elderly. International Encyclopedia of Public Health. 2008;254–60.

40. Health-Care Utilization as a Proxy in Disability Determination. Health-Care Utilization as a Proxy in Disability Determination. 2018 Apr 2;

41. Levesque JF, Harris MF, Russell G. Patient-centred access to health care: Conceptualising access at the interface of health systems and populations. Int J Equity Health [Internet]. 2013 Mar 11 [cited 2024 Mar 22];12(1):1–9. Available from: https://equityhealthj.biomedcentral.com/articles/10.1186/1475-9276-12-18

42. COREQ (COnsolidated criteria for REporting Qualitative research) Checklist.

43. Committee NHSR. The National Health Sciences Research Committee has general guidelines on health research. 2007;(December).

44. Rodríguez-Torres E, González-Pérez MM, Díaz-Pérez C. Barriers and facilitators to the participation of subjects in clinical trials: An overview of reviews. Contemp Clin Trials Commun [Internet]. 2021 Sep 1 [cited 2024 Dec 2];23:100829. Available from: https://pmc.ncbi.nlm.nih.gov/articles/PMC8358641/

45. National Institute for Health-Office of Research on Women’s Health. Review of the Literature: Primary Barriers and Facilitators to Participation in Clinical Research [Internet]. [cited 2024 Dec 2]. Available from: https://orwh.od.nih.gov/sites/orwh/files/docs/orwh_outreach_toolkit_litreview.pdf

46. Fogel DB. Factors associated with clinical trials that fail and opportunities for improving the likelihood of success: A review. Contemp Clin Trials Commun. 2018 Sep 1;11:156–64.

47. Mayson E, Ampt AJ, Shand AW, Ford JB. Intravenous iron: Barriers and facilitators to its use at nine maternity hospitals in New South Wales, Australia. Australian and New Zealand Journal of Obstetrics and Gynaecology. 2016;56(2):162–72.

48. Eke PC, Ossai EN, Eze II, Ogbonnaya LU. Exploring providers’ perceived barriers to utilisation of antenatal and delivery services in urban and rural communities of Ebonyi state, Nigeria: A qualitative study. PLoS One. 2021 May 1;16(5 May).

49. Uldbjerg CS, Schramm S, Kaducu FO, Ovuga E, Sodemann M. Perceived barriers to utilisation of antenatal care services in northern Uganda: A qualitative study. Sexual & Reproductive Healthcare. 2020 Mar 1;23:100464.

50. Sialubanje C, Massar K, Hamer DH, Ruiter RAC. Understanding the psychosocial and environmental factors and barriers affecting utilisation of maternal healthcare services in Kalomo, Zambia: a qualitative study. Health Educ Res. 2014;29(3):521–32.

51. Irwinda R, Wibowo N, Putri AS. Exploring the Determinants of Antenatal Care Services Uptake: A Qualitative Study among Women in a Rural Community in Northern Ghana. J Pregnancy. 2019;2019:1–6.

52. Abdiwali SA, Adesina OA, Fekadu GA, Geta TG. Barriers and facilitators to antenatal care services utilisation in Somaliland: a qualitative study. BMJ Open [Internet]. 2024 Nov 1 [cited 2024 Dec 2];14(11):e085073. Available from: https://bmjopen.bmj.com/content/14/11/e085073

53. Wilunda C, Scanagatta C, Putoto G, Montalbetti F, Segafredo G, Takahashi R, et al. Barriers to utilisation of antenatal care services in South Sudan: a qualitative study in Rumbek North County. Reprod Health [Internet]. 2017 May 22 [cited 2024 Dec 9];14(1):1–10. Available from: https://reproductive-health-journal.biomedcentral.com/articles/10.1186/s12978-017-0327-0

54. Malawi Multiple Indicator Cluster Survey Report | UNICEF Malawi [Internet]. [cited 2024 Dec 9]. Available from: https://www.unicef.org/malawi/reports/malawi-multiple-indicator-cluster-survey-report

55. Mweemba O, Zimba C, Chi BH, Chibwe KF, Dunda W, Freeborn K, et al. Contextualising men’s role and participation in PMTCT programmes in Malawi and Zambia: A hegemonic masculinity perspective. Glob Public Health [Internet]. 2022 Sep 2 [cited 2024 Dec 10];17(9):2081–94. Available from: https://www.tandfonline.com/doi/abs/10.1080/17441692.2021.1964559

56. Abdiwali SA, Adesina OA, Fekadu GA, Geta TG. Barriers and facilitators to antenatal care services utilisation in Somaliland: a qualitative study. BMJ Open [Internet]. 2024 Nov 1 [cited 2024 Dec 2];14(11):e085073. Available from: https://bmjopen.bmj.com/content/14/11/e085073

